# A circulating extracellular vesicle-bound fraction of cardiac troponin discriminates myocardial homeostasis and disease states

**DOI:** 10.64898/2026.03.16.26348488

**Authors:** Jona Benjamin Krohn, Dominika Bernath-Nagy, Yuetong Leona Ding, Melek Sukran Kalinyaprak, Gabriel Jakob Trauner, Chiara Hess, Felix Wiedmann, Constanze Schmidt, Hugo Katus, Norbert Frey, Florian Leuschner, Evangelos Giannitsis

## Abstract

**Background:** Various cardiovascular diseases are associated with transient troponin elevation, warranting further potentially invasive diagnostic measures to rule out myocardial ischemia. The origin of circulating cardiac troponins in the absence of overt myocardial necrosis remains unclear. Extracellular vesicles (EV) secreted by cardiomyocytes were found to contain cardiac troponins of unknown significance to date.

**Objectives:** We aim to investigate the presence of distinct distribution patterns of cardiac troponins in circulating EV and free plasma.

**Methods:** H9c2 cardiomyocytes exposed to hypoxia were investigated for troponin disintegration and vesicular troponin secretion. In a murine model of myocardial infarction and porcine model of atrial fibrillation, EV secretion into blood and EV-bound troponin fraction were studied. In two patient cohorts encompassing patients with myocardial infarction, tachyarrhythmia and healthy controls, EV- and non-EV-fraction troponin patterns were quantified.

**Results:** Hypoxic conditioning of cardiomyocytes enhanced EV-bound troponin T secretion. Minimal invasive myocardial infarction in mice caused pronounced systemic EV release. Induction of atrial fibrillation in pigs induced EV release and a shift in relative circulating troponin T compartmentalization. In patients presenting with tachyarrhythmia and myocardial infarction, elevated circulating EV concentrations were observed, concomitant with disease-specific relative EV-troponin fractional signatures in blood, concurrent with observations made in animal models. Circulating EV-troponin compartmental signatures could robustly discriminate myocardial infarction from tachyarrhythmia-induced myocardial injury, and further distinguish first diagnosis from recurrent tachyarrhythmia.

**Conclusions:** This study introduces the relative EV-troponin fraction as a novel biomarker in cardiovascular disease, improving diagnostic specificity for ischemic and non-ischemic myocardial disease entities.

## Introduction

Cardiovascular disease (CVD) with its potentially fatal sequelae myocardial infarction and ischemic stroke is the leading cause of morbidity and mortality worldwide (1). Highly sensitive cardiac troponin assays (hs-cTnT and hs-cTnI) constitute the diagnostic gold standard for transient or persistent myocardial ischemia (2), allowing for a timely detection of myocardial injury using validated diagnostic algorithms. The emergence of highly sensitive troponin assays in recent years, however, brought forth a novel challenge with regard to diagnostic specificity: a multitude of disease states beyond overt myocardial necrosis, such as tachyarrhythmias, sepsis, congestive heart failure or carditis, have been linked to often mild and/or transient cardiac troponin elevation in blood, following release kinetics inconsistent with those observed in myocardial ischemia (3). Clinical interpretation of elevated troponin levels in the context of cardiovascular disease are therefore complicated by the varying, oftentimes poorly understood pathophysiological mechanisms underlying time-dependent troponin release and degradation, particularly in the absence of clinically overt myocardial necrosis (4). Several hypotheses about cardiac troponin release from intact cardiomyocytes were stipulated in recent literature, postulating a transient increase in cell permeability or cell membrane “blebbing” under pathological conditions (5).

Extracellular vesicles (EV) are membrane-bound nanoparticles released by a multitude of different cell types, their functions spanning autocrine as well as paracrine cell-cell communication through specific delivery of proteins or RNA, as well as interaction of cells with the extracellular matrix (6). EV carry a specific molecular cargo reflecting on the current state of their cell of origin (7). Emerging evidence on the diagnostic potential of EV in cardiovascular disease has linked myocardial hypoxic stress to an increase in quantitative EV release (8), and numerous clinical studies have focused on the biomarker potential of circulating EV and EV-enclosed molecular targets in different cardiovascular pathologies such as coronary artery disease and myocardial infarction (9–12), arrhythmias (13, 14) or heart failure (15, 16). A promising link between the abundance of distinct EV populations in circulation and cardiovascular prognosis holds immense diagnostic potential, warranting further exploration (17). Few studies to date have identified the presence of cardiac troponins in cardiomyocyte-derived circulating EV (18, 19), while their diagnostic role, functional implications and contribution to total blood troponin levels in health and disease remain nebulous.

This study investigates the effect of myocardial strain and ischemia on quantitative EV release and EV enrichment with cardiac troponins T and I, which is postulated to hold the potential to discriminate temporary myocardial strain from irreversible myocardial necrosis. We hypothesize that particularly the early stages of the myocardial stress response may be characterized by expeditious EV secretion and specific enrichment with structural troponins of distinct fractional composition, improving diagnostic specificity with regard to ischemic and non-ischemic myocardial disease entities.

## Methods

### Patient recruitment

Patients were recruited from the Chest Pain Unit (emergency department) at University Hospital Heidelberg, Germany, following informed written consent. Individuals with non-ST elevation myocardial infarction (NSTEMI) requiring percutaneous coronary intervention and atrial fibrillation or atrial flutter (AF) were diagnosed according to current guidelines. Healthy control patients were identified based on unsuspicious or nonspecific symptoms and guideline-conform rule-out of myocardial disease through appropriate diagnostic testing. Routine phlebotomy as well as further analyses were performed in consensus with the institutional review board at the Faculty of Medicine of Heidelberg University (IRB Approval #S-351/2015) in accordance with the Declaration of Helsinki in its latest amendment.

### PB-EV isolation

Citrate-supplemented venous blood samples were obtained via routine phlebotomy and processed within 120 minutes, avoiding agitation. Subsequent EV isolation was essentially conducted as described previously (20), with some slight modifications. In brief, whole blood samples were diluted in phosphate buffered saline (PBS) to reduce viscosity and Histopaque was used to promote separation. Plasma was separated through density gradient centrifugation. Using incremental centrifugation steps starting at 2500 times g for 15 mins to remove large cellular debris with a final ultracentrifugation at 10^5^ times g for 1 hour using an XPN-80 ultracentrifuge with a TLA 50.2 Ti fixed-angle rotor (Beckman Coulter) for optimal pellet visualization, EV were pelleted and carefully separated from the EV-depleted supernatant. EV pellets were resuspended and homogenized instantly in a solution of choice depending on downstream analysis and analyzed immediately or stored at -80°C until further use.

### Nanoparticle tracking analysis

Pelleted EV were resuspended in PBS and diluted accordingly to obtain a suitable working solution resulting in a particle density of 10-100 EV per frame upon nanoparticle tracking analysis (NanoSight NS300 using Nanosight NTA 3.2 Software, Malvern Panalytical). Diluted EV suspensions were analyzed with a constant camera setting of 9 and detection threshold of 2 across all samples, and nanoparticle concentration and size distribution was averaged from three recorded 30-second videos.

### Fractional cTnT and cTnI quantification

EV-cTnT and -cTnI levels were quantified by resuspending separated EV pellets in radioimmunoprecipitation assay-based EV-membrane lysis buffer (Abcam). EV-lysates were diluted with PBS and hs-cTnT and hs-cTnI were measured by high-sensitive assays used in everyday clinical practice (Cobas E 411, Roche Diagnostics; ATELLICA hs-cTnI Assay, Siemens Healthcare Diagnostics). Non-EV-cTn quantified from the EV-depleted plasma from each sample as well as cTnT/cTnI quantified from simultaneous routine blood draw were measured using the same assays.

### Immunoblotting for specific EV markers

Enrichment of EV-specific markers in EV lysates were demonstrated by immunoblot in accordance with current MISEV recommendations (21). Briefly, EV-pellets resuspended in radioimmunoprecipitation assay-based EV-membrane lysis buffer (Abcam) and appropriate corresponding volumes of EV-depleted plasma were sonicated and total protein concentration of the samples were measured by BCA assay (Thermo Fisher Scientific). Proteins were denatured at 95°C and equal amounts were loaded on to 4-12% polyacrylamide gels containing sodium dodecyl sulfate and separated through electrophoresis at constant voltage. Separated proteins were blotted onto polyvinylidene difluoride membranes and nonspecific binding sites were blocked with bovine serum albumin. Membranes were incubated with primary antibodies (anti-CD9, Cell Signaling Technology; anti-Annexin 5, Cell Signaling Technology; anti-TSG101, Abcam) diluted accordingly at 4°C overnight under agitation. After washing, membranes were incubated with horse radish peroxidase-conjugated species-specific secondary antibodies (Abcam) diluted accordingly for 1 hour at RT. After washing, bands were detected using the SuperSignal West Femto Maximum Sensitivity Substrate (Thermo Fisher) on a Fusion FX imaging system (Vilber Lourmat).

### cTnT Immunoprecipitation and immunoblot from EV and EV-depleted plasma

EV pellets were lysed in a solubilization buffer containing Triton X-100 (Merck) and the corresponding volume of EV-depleted plasma was diluted 1:1 in the same buffer. Biotinylated M11-7 anti-cTnT antibody kindly provided by Roche Diagnostics was diluted to a concentration of 5 μg/ml and added to EV isolates and the EV-depleted plasma. Streptavidin Dynabeads (Invitrogen) were used for cTnT pulldown according to manufacturer’s instructions. Positive control was prepared using recombinant human cTnT (Hytest) diluted to 500 pg/ml and processed in parallel. Following overnight incubation at constant agitation, cTnT was eluted from the beads through denaturation.

Eluted denatured cTnT was loaded on to 4-12% polyacrylamide gels containing sodium dodecyl sulfate and proteins were separated as described previously. Gels were blotted onto polyvinylidene difluoride membranes and non-specific binding sites were blocked with bovine serum albumine. Membranes were incubated with unconjugated M11-7 anti-cTnT antibody (Roche Diagnostics) diluted accordingly overnight under constant agitation. After washing, membranes were incubated with horse radish peroxidase-conjugated secondary antibody (Abcam) for 1 hour at room temperature. Bands were detected using the SuperSignal West Femto Maximum Sensitivity Substrate (Thermo Fisher) on a Fusion FX imaging system (Vilber Lourmat).

### Differentiation and hypoxic conditioning of rat H9c2 cardiomyoblasts

Rat cardiomyoblast cell line H9c2 was cultured in Dulbecco’s modified Eagle medium (DMEM/F-12, Gibco) containing 10% fetal bovine serum and 1% penicillin/streptomycin, and cells were differentiated to a cardiomyocyte-like phenotype in the presence of all-trans retinoic acid as described elsewhere (22). Following serum starvation at 1% EV-depleted fetal bovine serum, cells were exposed to 1.5% atmospheric O_2_ for 6, 12 and 24 hours and control cells were incubated at normoxia. Following incubation, cell supernatant was removed and EV were isolated by specific precipitation (Total Exosome Isolation Reagent from cell culture media, Thermo Fisher Scientific). Cells were washed and lysed, and cTnT was detected by immunoblotting as described previously. EV-cTnT was detected using immunoprecipitation followed by immunoblotting as described above.

### H9c2 transfection and immunofluorescence

H9c2 cells were transfected with either His-tagged TNNT2 plasmid or Luciferase control plasmid (Vectorbuilder) using Lipofectamine 3000 according to manufacturer’s instructions (Thermo Fisher Scientific) before differentiation in starvation medium as described above. Cells were exposed to 1.5% atmospheric O2 for 12 hours and compared to normoxic control. Cells were subsequently washed and fixed with 4% paraformaldehyde (PFA), cell membranes were permeabilized with 0.1% Triton X-100 (Merck) and nonspecific binding sites were blocked with bovine serum albumin. Cells were incubated with appropriate primary antibodies (Abcam) overnight at 4°C. After washing, fixed cells were incubated with appropriate Alexa Fluor-conjugated secondary antibodies (Abcam) for 1 hour at RT. After washing, cell nuclei were stained with 4′,6-Diamidino-2-phenylindol. Imaging was performed on a Leica SP8 confocal microscope using the Z-scan technique.

### Live/dead cell assay

Differentiated H9c2 cells were detached with 1x non-enzymatic dissociation reagent (Sigma Aldrich) at 37°C for 30 mins and quenched with equal volumes of cell culture medium. Following complete removal of cells from culture surface, cell suspensions were centrifuged at 300 times g for 10 mins at 4°C, and pelleted cells were resuspended in fluorescence-activated cell sorting (FACS) buffer containing Hank’s balanced salt solution with 1% fetal calf serum and 2mM ethylene diamine tetraacetate. 10^6^ cells per sample were stained with the Live/Dead Cell Stain Kit (Thermo Fisher Scientific) according to manufacturer’s instructions. Following centrifugation and washing, flow cytometry was performed on a FACSVerse multicolor flow cytometer (BD Biosciences), and quantitative analysis was done using FlowJo software.

### Murine minimal-invasive myocardial infarction model (MiMi)

All animal experiments were conducted in accordance with the German Law on the Protection of Animals and the ARRIVE guidelines in its latest amendment following approval by the institutional review board at Regierungspräsidium Karlsruhe (IRB# G-127/19). Wild type C57BL6/J mice aged 18-21 weeks underwent an induced myocardial infarction as described by Sicklinger et al. (23), whereby ligation of the left anterior descending artery (LAD) was achieved through a minimal invasive approach. Mice undergoing a sham procedure without LAD coagulation served as control. Four hours following MiMi or sham, animals were euthanized, and blood and hearts were harvested. Whole heart sections were cut into 10 μm slices and stained with Masson Trichrome to visualize the infarcted area. cTnT was measured using the high sensitive assay used for all human samples (Cobas E 411, Roche Diagnostics). EV were isolated using an EV precipitation kit (Thermo Fisher Scientific) according to manufacturer’s instructions, and quantification of EV concentrations was achieved through nanoparticle tracking analysis as described previously.

### Porcine model of pacemaker-induced tachyarrhythmia

All animal experiments were conducted in accordance with the German Law on the Protection of Animals and the ARRIVE guidelines in its latest amendment, following institutional review board approval (Regierungspräsidium Karlsruhe, IRB# G-131/22). Procedures were performed as described in detail in a previous publication by Paasche, Wiedmann et al (24). In brief, adult German landrace pigs were anesthetized and treated with a dual-lead pacemaker and central venous catheter (CVC) implantation on day 0 of the experiment. Blood was drawn from the CVC, and atrial fibrillation was induced and sustained by pacemaker-induced continuous loop atrial burst stimulation. Persistent AF was confirmed through electrocardiogram on day 3, and after a total duration of 5 days under tachyarrhythmia, blood was drawn from the CVC, and plasma EV were isolated through incremental centrifugation steps and a subsequent ultracentrifugation as described above. Quantification of EV concentrations was achieved through nanoparticle tracking analysis as described previously. Fractional cTnT quantification from native plasma, EV isolates and EV-depleted plasma samples was implemented using gold standard high-sensitive assay (Cobas E 411, Roche Diagnostics) as described above.

### Statistics

All statistical analyses were performed using GraphPad Prism version 10.2.3. Sample size n is generally stated as the number of biological replicates (patients, animals) to reflect biological variance, while a sufficient number of technical replicates was conducted as feasible to account for experiment-associated noise. For the H9c2 cell line, sample size n corresponds to the number of independent experiments. Shapiro-Wilk test was applied to test for normality where appropriate. For normally distributed parameters comparing three groups, one-way analysis of variance (ANOVA) with Bonferroni post-hoc test for multiple comparisons was used. For non-normally distributed parameters, Kruskal-Wallis test with Dunn’s multiple comparison test was applied. Data is presented as mean±standard deviation, and error bars in graphs represent standard deviation, unless otherwise specified. A p-value of <0.05 was considered statistically significant. For correlation analyses in Fig. 5, linear regression was performed, and Pearson’s R^2^ was calculated. To test for significance of correlation, a t-test was performed assuming H0: β = 0 and α = 0.05.

## Results

### Hypoxic stress prompts structural troponin disintegration in cardiomyocytes with subsequent enrichment in secreted EV

To explore the hypothesis of an EV-mediated troponin release mechanism in response to stress, the rat cardiomyoblast cell line H9c2 was differentiated into a cardiomyocyte-like phenotype and exposed to incremental hypoxia (Fig. 1A). Compared to baseline, conditioning with 1.5% atmospheric O_2_ at 6 and 24 hours resulted in an upregulation of intracellular cardiac troponin T (cTnT) concomitant with a pronounced enrichment in secreted EV after 24 hours of hypoxia (Fig. 1B+C, Suppl. Fig. S1C). To rule out apoptotic cell disintegration as a potential contributing factor, cell viability was found unaltered compared to normoxic conditions (Suppl. Fig. S1B). To visualize structural troponin T fragmentation under hypoxic stress, differentiated H9c2 cells were transfected with His-tagged Tnnt2 or Luciferase control (Fig. 1D, Suppl. Fig. S1A) to ensure specific detection under hypoxia. After 12 hours of hypoxic conditioning of H9c2 cardiomyocytes, intracellular bodies containing His-tagged cTnT and cysteine protease calpain were detected by immunofluorescence (Fig. 1E). Calpain has previously been identified as a key enzyme in the constitutive and disease-specific degradation of cardiac troponins T and I in cardiomyocytes (25, 26). These findings may indicate a distinct pathway of structural troponin disintegration and sequestration into secretory EV in response to reversible myocardial injury, a long-disputed phenomenon that may account for temporary systemic troponin elevation in the absence of myocardial necrosis.

**Fig. 1:**
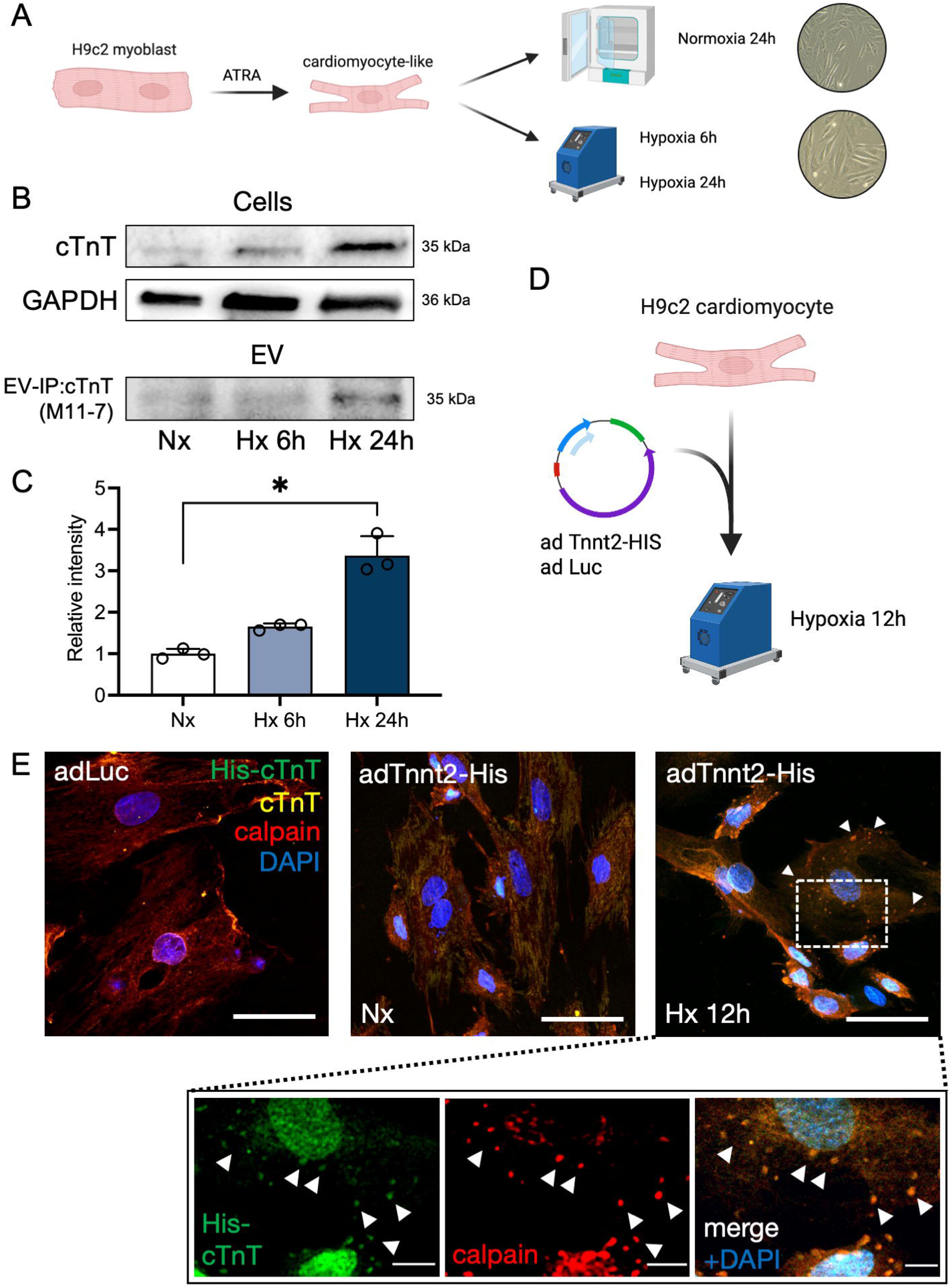
Hypoxia triggers intracellular troponin T disintegration in cardiomyocytes and vesicular cTnT enrichment in vitro. **A** Rat H9c2 myoblasts differentiated into a cardiomyocyte-like phenotype in the presence of ATRA are exposed to incremental periods of hypoxia at 1% atmospheric O_2_ (*created in BioRender, License #TL29CBWSPH*). **B** Hypoxic conditions trigger cTnT expression in H9c2 cells (*top*) and incremental enrichment in secreted EV detected by cTnT immunoprecipitation (*bottom*) using Roche’s M11-7 biotinylated antibody. **C** Quantification of relative band intensity of EV-IP:cTnT after 6 and 24 hours of hypoxia normalized to normoxia control. **D** Serum-starved H9c2 cardiomyocytes are transfected with either His-tagged Tnnt2 or luciferase plasmid (*created in BioRender, License #NJ29CC7PUC*). **E** Transfection of Tnnt2-His or luciferase control plasmid to differentiated H9c2 cardiomyocyte-like cells reveals progressive granulation of structural cTnT and co-localization with calpain (arrowheads) indicating lysosomal fragmentation under hypoxic stress. *Inlay*: co-localization of His-cTnT with calpain in cytoplasm. *63x magnification, scale bar 5* μ*m (2* μ*m for inlay)*. *ATRA = all-trans retinoic acid, cTnT = cardiac troponin T, Nx = normoxia, Hx = hypoxia. n≥3 independent experiments*.

### Animal models of irreversible myocardial necrosis and reversible myocardial strain recapitulate systemic EV release and distinct cTnT compartmentalization

To verify our in vitro findings on the release of troponin-enriched EV in response to myocardial stress at organismal level, we exploited a previously established murine model of minimal-invasive myocardial infarction (MiMi, Fig. 2A) (23). This minimal-invasive, non-surgical approach is ideal for the study of systemic EV secretion in response to myocardial ischemia, as it minimizes procedure-related systemic troponin and EV release. To observe the early effects of electrical coagulation of the left anterior descending (LAD) coronary artery, mice are sacrificed after 4 hours and blood and whole-heart tissue are analyzed. Prominent thinning of the anterior-wall myocardium not observed in sham controls is appreciated as an early sign of ischemia (Fig. 2B), concomitant with an increase in total plasma cTnT (14120±9319 pg/ml in MiMi vs. 290±170 pg/ml in sham, p=0.06) (Fig. 2C), confirming technical success of the procedure. As early as 4 hours post MiMi, a marked ∼4-fold rise in circulating EV concentration compared to sham is noted (2.8x10^11^±1.3x10^11^/ml in MiMi vs. 7.2x10^10^±2.3x10^10^/ml in sham, p=0.0003) (Fig. 2C+D), with typical EV size distribution upon nanoparticle tracking analysis (148±7 nm in MiMi vs. 153±22 nm in sham, p=0.81) (Fig. 2E). These results suggest plasma-derived EV as an early-response marker for myocardial ischemia with potential additive diagnostic value.

**Fig. 2:**
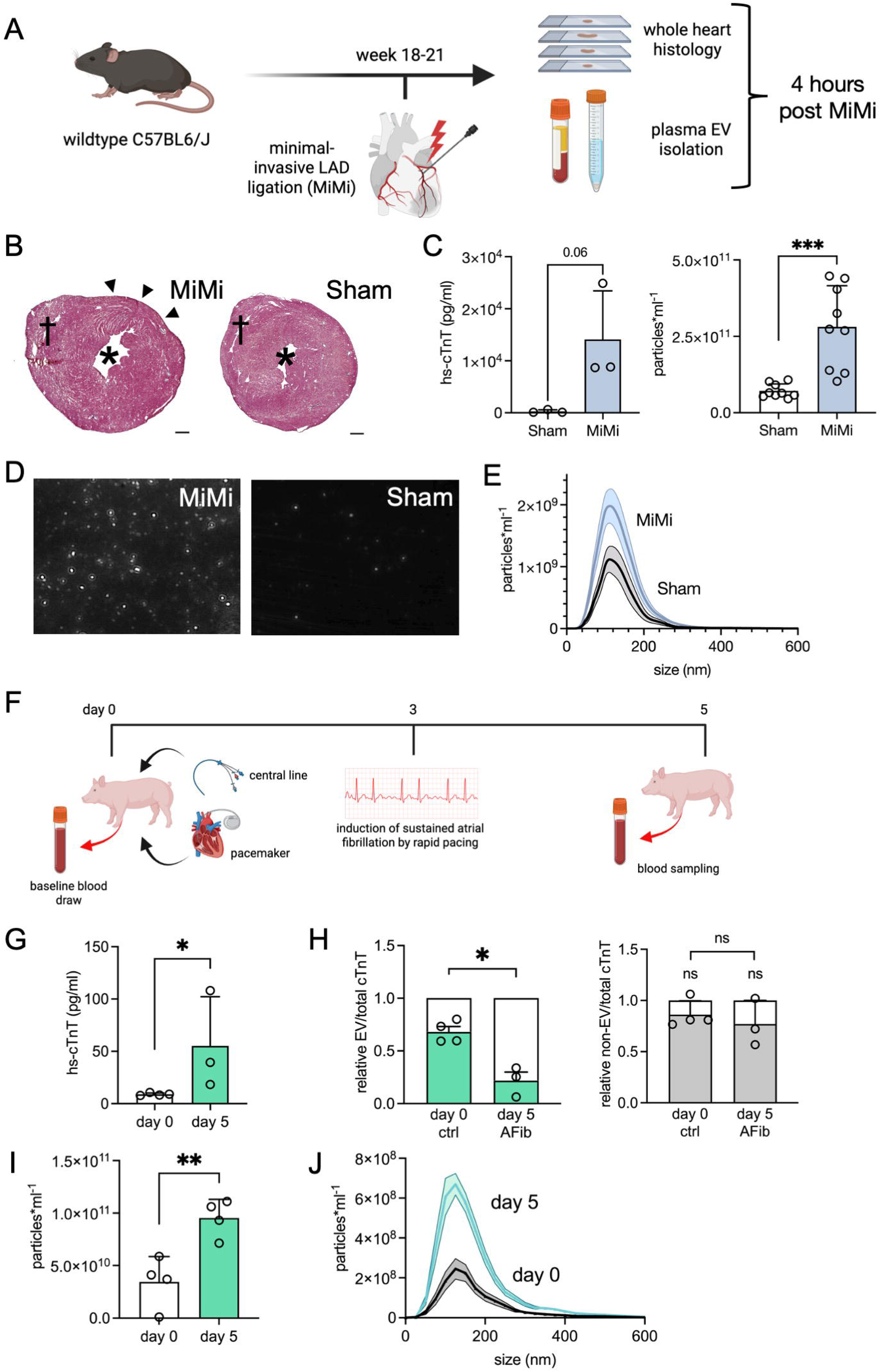
A murine model of myocardial infarction and porcine model of myocardial strain reproduce systemic EV release and EV-cTnT compartmentalization. **A** Wildtype mice are exposed to either minimal invasive LAD ligation (MiMi; n=3) or sham intervention (n=3). After 4 hours, plasma is obtained for EV isolation and whole hearts are harvested for histology (*created in BioRender, License #SF29CCHRAW*). **B** Masson Trichrome staining of whole heart sections reveals early signs of ischemia in the anterior segments following LAD coagulation, while no signs of ischemia are shown in sham mice. *\* left ventricle ^†^ right ventricle.* C Total plasma cTnT is elevated in MiMi mice compared to sham (*left*) consecutive to increased systemic EV release (*right*). **D** Representative NTA images of plasma EV isolates demonstrate a marked increase in circulatory EV 4 hours after MiMi compared to sham. **E** Size distribution of tracked EV from plasma of MiMi and sham mice showing no difference in EV size. **F** Induction of rapid pacing-induced tachyarrhythmia in pigs (n=4) (*created in BioRender, License #GE29CGY59H*). **G** Total plasma cTnT after tachyarrhythmia induction. **H** Relative EV-bound cTnT (*left*) and cTnT in EV-depleted plasma (*right*) normalized to total plasma cTnT at days 0 and 5. **I** Plasma EV concentration and **J** EV size distribution at days 0 and 5 quantified by NTA. *NTA = Nanoparticle tracking analysis. *p<0.05, **p<0.01, ns=not significant*.

To investigate the dynamics of systemic EV release in response to temporary myocardial strain, a porcine model of atrial fibrillation (AF) is used. Herein, adult landrace pigs receive a baseline blood draw for EV isolation to serve as intrinsic control, followed by dual-chamber pacemaker implantation and induction of AF by atrial burst pacing (24) (Fig. 2F). After 72 hours of sustained tachyarrhythmia, blood sampling reveals consecutively elevated plasma cTnT levels (55.2±46.9 pg/ml under AF vs. 8.9±1.3 pg/ml at baseline, p=0.048) (Fig. 2G). Consistent with the murine model of myocardial ischemia, tachyarrhythmia-induced strain associates with a marked ∼3-fold increase in total circulating EV levels (9.5x10^10^±1.8x10^10^/ml under AF vs. 3.4x10^10^±2.4x10^10^/ml at baseline, p=0.008) (Fig. 2I) of typical size distribution (178±20 nm under AF vs. 178±17 nm at baseline, p=0.98) (Fig. 2J). An in-depth analysis of the distribution of cTnT in EV relative to free plasma reveals a shift of total circulating cTnT towards the free plasma compartment under sustained AF (Fig. 2H), which may indicate either progressive cTnT liberation from apoptotic cardiomyocytes under prolonged strain, or a progressive release of cTnT from circulating EV through EV degradation. No difference is noted between cTnT from EV-depleted plasma (termed ‘non-EV cTnT’) and total plasma cTnT, suggesting that both cTnT levels measure the same free plasma compartment.

Our findings purport that in both animal models of myocardial ischemia and arrhythmia-induced strain, the myocardial stress response associates with accelerated circulating EV release. Further, sustained myocardial strain induces a shift of cTnT from EV to free-serum compartments in plasma, prompting a distinct EV/free plasma cTnT compartmental signature.

### Quantitative circulating EV levels and relative plasma EV-cTnT compartmentalization reveal distinct EV-cTnT signatures in health and disease states

Present *in vitro* and *in vivo* data suggest a mechanistic connection between myocardial hypoxic strain and ischemia with quantitative EV release and specific blood cTnT compartmentalization. To translate the concept of a differential cTnT enrichment in circulating cardiomyocyte-derived EV, two independent patient cohorts, each encompassing three groups, were recruited from the emergency department (Chest Pain Unit) at University Hospital Heidelberg: Patients presenting with acute myocardial strain (tachyarrhythmic atrial fibrillation or flutter, AF), acute myocardial ischemia (non-ST elevation myocardial infarction, NSTEMI) requiring percutaneous coronary intervention, or non-specific symptoms not suspicious of myocardial disease (control). From all patients, blood was drawn with routine bloodwork upon presentation, and EV isolates and EV-depleted plasma compartments were analyzed separately. In the pilot cohort, NSTEMI and AF patients were older on average and exhibited a reduced renal function compared to the control group, while no difference was found in body mass index, sex distribution, blood pressure upon presentation, or time from symptom onset (Table 1). Naturally, plasma cTnT levels were higher in the NSTEMI and AF groups, as was heart rate in patients presenting with symptomatic tachyarrhythmia. In the validation cohort, similar trends were observed (Table 2).

In alignment with animal data, global circulating EV levels were markedly elevated in the setting of tachyarrhythmic strain (AF; 6.3x10^10^±4.1x10^10^/ml, p<0.0001 vs. control), and even more pronounced in myocardial ischemia (NSTEMI; 9.9x10^10^±8.3x10^10^/ml, p<0.0001 vs. control and AF, respectively) compared to control (3.4x10^10^±2,3x10^10^/ml; Fig. 3A), while average EV size did not differ significantly between the groups (169.8±47.7 nm for AF, 181.2±23.0 nm for NSTEMI, 188.3±47.7 nm for control; p=0.41 AF vs. NSTEMI, p= 0.21 control vs. NSTEMI, p>0.99 control vs. AF; Fig. 3B). Adequate separation of EV from plasma compartments was verified by immunoblotting for EV-enriched protein targets CD9, annexin 5 and TSG101 (Fig. 3C, Suppl. Fig. S2A-C). Immunoprecipitation of cTnT contained in EV isolates and corresponding EV-depleted plasma volumes from a patient presenting with NSTEMI and a healthy control patient revealed different relative cTnT content enclosed within EV and in free plasma between the two patients (Fig. 3D, Suppl. Fig. S3), warranting further quantitative analysis.

**Fig. 3:**
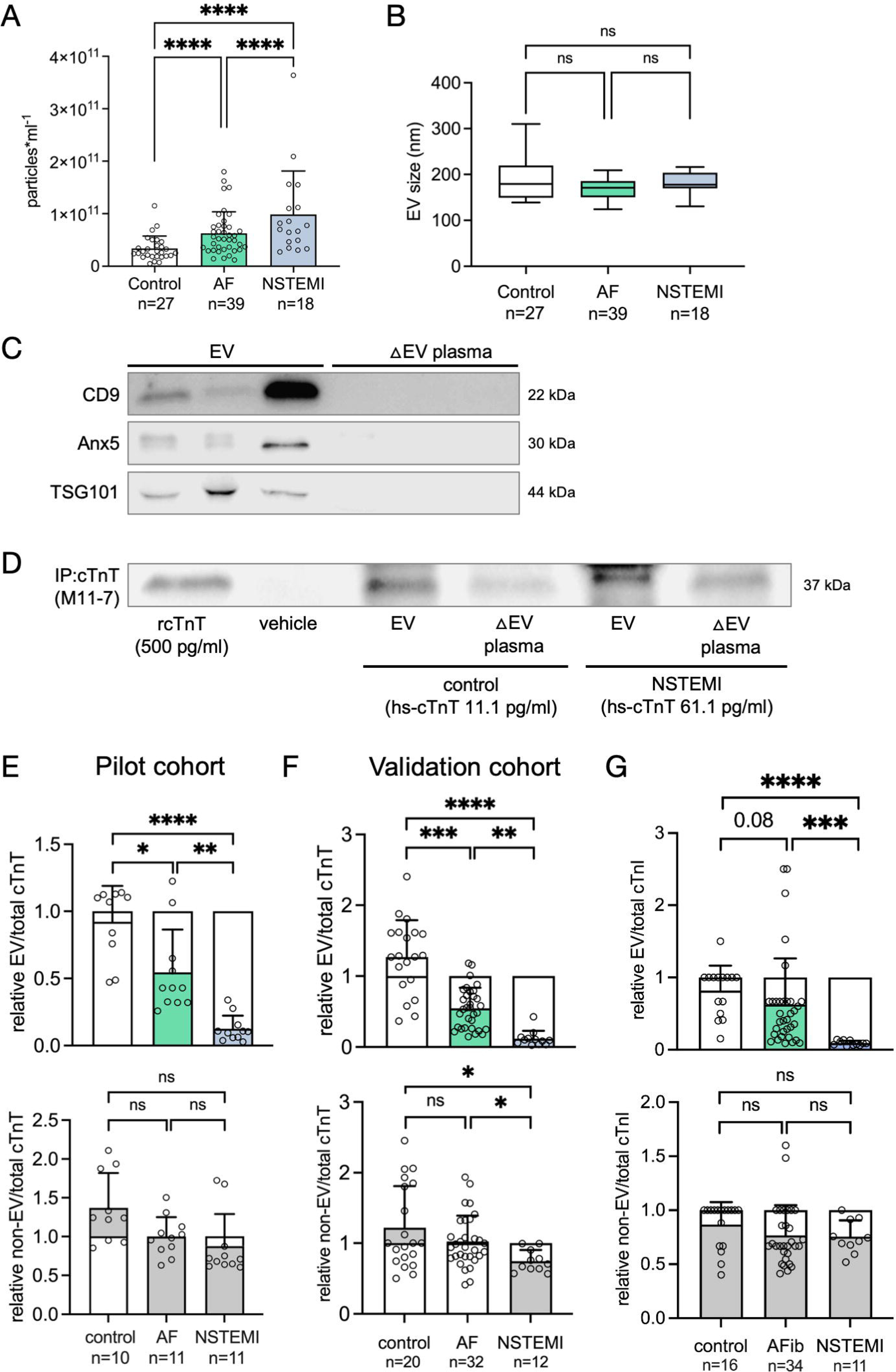
Plasma-derived EV from patients with myocardial ischemia (NSTEMI) and tachyarrhythmia-induced myocardial strain (AF) exhibit distinct EV-bound cardiac troponin signatures. **A** EV concentration from plasma isolates is incrementally increased in the setting of atrial fibrillation or flutter (Afib/Aflut) and NSTEMI compared to healthy controls. **B** No significant difference in EV size is appreciated between experimental groups. **C** Isolation of EV is verified by immunoblotting for EV-enriched markers CD9, annexin 5 (Anx5) and TSG101 in EV isolates and EV-depleted plasma samples. n=3 biological replicates. **D** High-sensitive cTnT immunoprecipitation of EV isolates and EV-depleted plasma of a control and NSTEMI patient shows differential cTnT enrichment across EV and non-EV compartments. **E** Relative enrichment of EV-cTnT (*top*) and non-EV-cTnT (*bottom*) normalized to total plasma cTnT in atrial fibrillation (AF), NSTEMI and healthy controls reveals distinct cTnT compartmentalization dependent on myocardial pathophysiology in both a pilot cohort and an independent validation cohort (**F**). **G** Relative enrichment of EV-cardiac troponin I (cTnI; *top*) and non-EV-cTnI (*bottom*) normalized to total plasma cTnI in AF, NSTEMI and healthy controls reveals disease-specific cTnI blood compartmentalization. *\*p<0.05, **p<0.01, ***p<0.001, ****p<0.0001, ns=not significant*.

Following separation of plasma-derived EV and EV-depleted plasma compartments, cTnT content was quantified separately and normalized to free-plasma cTnT determined as part of clinical routine, all using the same high-sensitive assay. In both pilot and validation cohorts, normalized EV-bound cTnT as well as EV-depleted plasma cTnT in the healthy control group did not differ from routine free-plasma cTnT levels (Fig. 3E+F), raising the hypotheses that 1. in cardiac homeostasis, total circulating cTnT is distributed in equal parts in EV and free-plasma compartments, and 2. routine blood cTnT levels only reflect free-plasma cTnT, whereas the EV-bound cTnT fraction largely evades current diagnostic standard. In the setting of myocardial strain secondary to tachyarrhythmia, EV-bound cTnT fraction consistently accounts for 54.6±31.8% (p=0.02 vs. control) and 54.7±28.8% (p=0.0004 vs. control) of routine free-plasma cTnT in the pilot and validation cohort, respectively (Fig. 3E+F). Compared to healthy control, these data suggest a significant compartmental shift of circulating cTnT favoring the free-plasma fraction under sustained myocardial strain. In myocardial ischemia, an even more pronounced redistribution of cTnT favoring the free-plasma compartment is observed with relative EV-bound cTnT making up a mere 12.5±9.9% (p=0.0011 vs. AF; p<0.0001 vs. control) and 11.9±10.9% (p=0.001 vs. AF; p<0.0001 vs. control) of routine free-plasma cTnT across both patient cohorts (Fig. 3E+F).

Samples in the validation cohort were further evaluated for cardiac troponin I (cTnI) content across blood compartments. While in the healthy control group and in myocardial ischemia, similar distribution of cTnI compared to cTnT was found in plasma EV relative to routine plasma cTnI (82.0±34.4% in healthy controls, 9.3±3.2% in NSTEMI patients; p<0.0001), in tachyarrhythmia-induced myocardial strain, cTnI content in EV was more heterogeneous and did not differ significantly to control (62.5±63.8%, p=0.08; Fig. 3G). Overall, our patient data from two independent cohorts reveal distinct cTnT and cTnI compartmental signatures potentially related to disease mechanism-specific release pathways of cardiac troponins into circulation.

### Cardiac troponins enclosed within the circulating EV fraction may improve diagnostic specificity in tachyarrhythmia-induced myocardial strain

Present evidence on the relative distribution of circulating cardiac troponins within blood-derived EV and free-plasma compartments suggest specific compartmental signatures depending on the underlying myocardial pathophysiology, pointing towards specific troponin release mechanisms from diseased cardiomyocytes. We further investigated whether cardiac troponin compartmental signatures may improve diagnostic specificity within the heterogeneous myocardial strain cohort, often associated with blood troponin dynamics holistically termed “type II myocardial infarction” or “myocardial infarction with non-occlusive coronary arteries” (MINOCA) in clinical routine. In two subgroup analyses, the AF cohort was further divided into patients with firstly diagnosed AF and recurrent episode of AF upon presentation, as well as patients with tachycardic AF (with a heart rate >100/min) and normofrequent AF (with a heart rate <100/min) upon presentation to the emergency department. No significant difference was appreciated in systemic EV concentration between fist-diagnosis and AF recurrence (7.5x10^10^±4.3x10^10^/ml vs. 6.3x10^10^±3.6x10^10^/ml; p>0.99), nor between normofrequent and tachycardic AF (6.6x10^10^±4.6x10^10^/ml vs. 6.9x10^10^±3.5x10^10^/ml; p>0.99) (Fig. 4A+B). Concurrent with previous data, average EV size was found to be similar in all experimental subgroups.

**Fig. 4:**
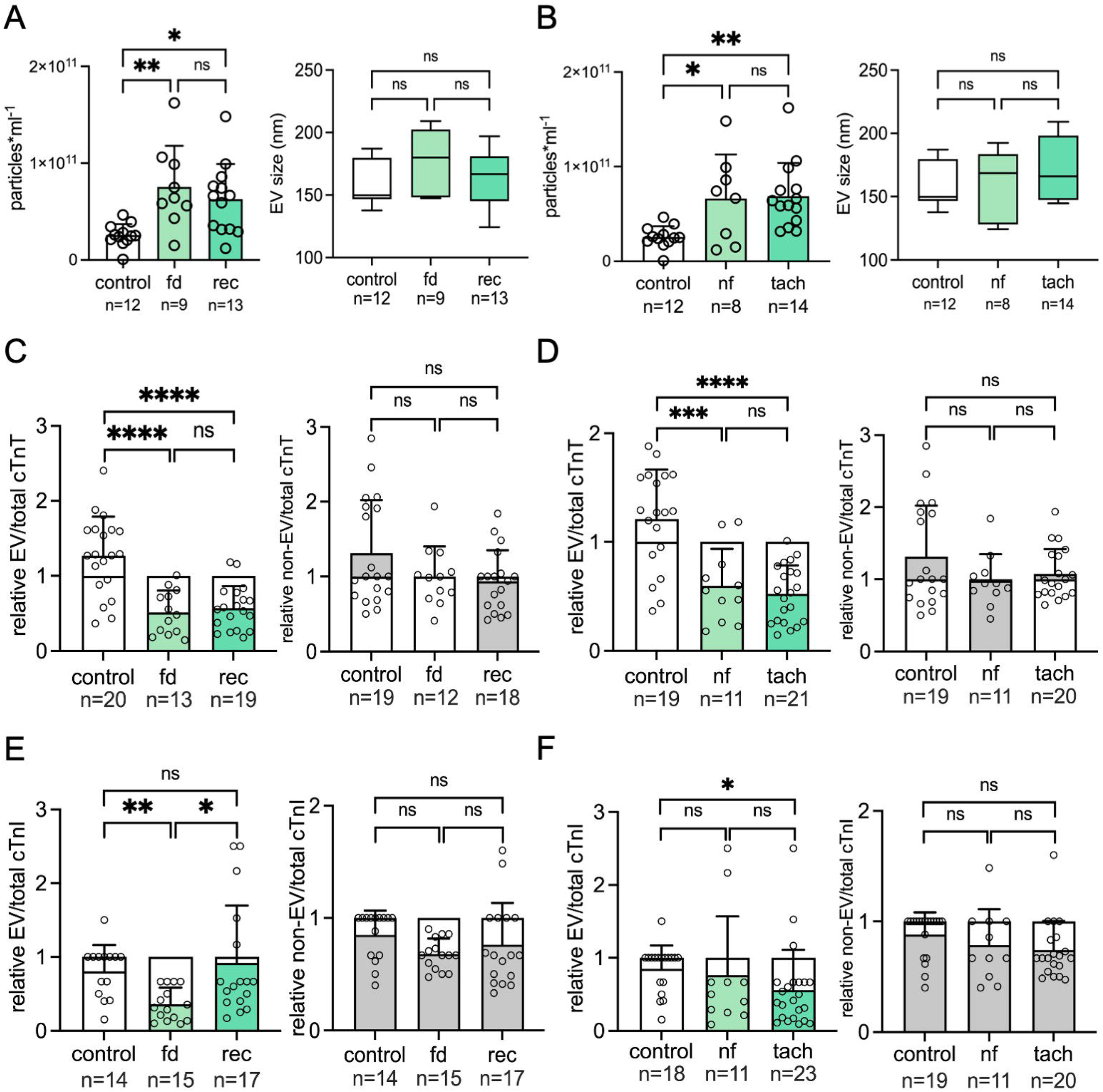
Stratification of the tachyarrhythmia-induced myocardial strain (AF) cohort in terms of quantitative EV release, EV size and EV-bound troponin T/I signatures. **A** Plasma EV concentration and EV size in firstly diagnosed (fd) vs. recurrent (rec) atrial fibrillation (AF) and **B** in normofrequent (nf) vs. tachyarrhythmic (tach) AF, respectively. **C** Relative enrichment of EV-cTnT (*left*) and non-EV-cTnT (*right*) normalized to total plasma cTnT in first diagnosis (fd) vs. recurrent (rec) AF, and **D** in normofrequent (nf) vs. tachyarrhythmic (tach) AF, respectively. **E** Relative enrichment of EV-cTnI (*left*) and non-EV-cTnI (*right*) normalized to total plasma cTnI in first diagnosis (fd) vs. recurrent (rec) AF, and **F** in normofrequent (nf) vs. tachyarrhythmic (tach) AF, respectively.

While relative EV-cTnT did not differ substantially between firstly diagnosed and recurrent AF (51.3±29.2% vs. 57.1±29.2%, p>0.99; Fig. 4C), nor between normofrequent and tachycardic AF (59.5±34.1% vs. 52.2±26.3%, p>0.99; Fig. 4D), relative EV-bound cTnI was found to discriminate firstly diagnosed AF (36.0±22.4%) from AF recurrence (92.1±77.5%, p=0.02 vs. first diagnosis, p>0.99 vs. control; Fig. 4E). Interestingly, the previously observed shift of cTnI in AF towards the free-plasma compartment could only be reproduced in firstly diagnosed AF, whereas in a recurrent AF episode, cTnI compartmental signatures were largely indistinguishable from healthy controls, potentially reflecting myocardial remodeling in response to persistent and/or recurrent AF. Based on heart rate, no distinction could be made with relative EV-bound cTnI as discriminator (76.5±80.4% for normofrequent, 55.9±55.0% for tachyarrhythmic patients, p>0.99; Fig. 4F). Furthermore, in accordance with present data, neither relative cTnT nor cTnI from EV-depleted plasma samples differed from routine free-plasma cTnT/cTnI levels throughout all experimental groups.

### Correlation of free-serum with EV-bound cardiac troponin hints at distinct release kinetics in myocardial strain and ischemia states

To further the hypothesis of distinct disease-specific troponin release mechanisms resulting in differential troponin compartmentalization patterns, we aimed to correlate free-plasma with EV-bound cTnT and cTnI in the investigated myocardial disease cohorts. In the context of overt myocardial ischemia, incipient myocardial strain and subsequent apoptosis with structural disintegration of the cardiomyocyte hypothetically leads to a proportional increase of both EV-bound and free-plasma cTnT and cTnI. Concurrent with this theory, the observed elevation in global circulating EV and routine plasma cTnT/cTnI associates with a positive correlation of EV-bound with free-plasma cTnT (R^2^ 0.67, p<0.0001; Fig. 5A) and cTnI (R^2^ 0.81, p<0.0001; Fig. 5B) in our ischemia cohort. This result may indicate an EV-bound troponin release largely driven by cellular disintegration. In tachyarrhythmia-induced myocardial strain, however, EV-bound cTnT does not correlate with free-plasma cTnT (R^2^ 0.0008, p=0.98; Fig. 5C), and while a correlation of EV-bound with free-plasma cTnI can be established (R^2^ 0.48, p<0.0001; Fig. 5D), correlation is less stringent compared to the ischemia cohort. These observations suggest a competing mechanism of EV-troponin enrichment and/or troponin release into free plasma beyond cardiomyocyte structural disintegration in the setting of temporary myocardial strain.

**Fig. 5:**
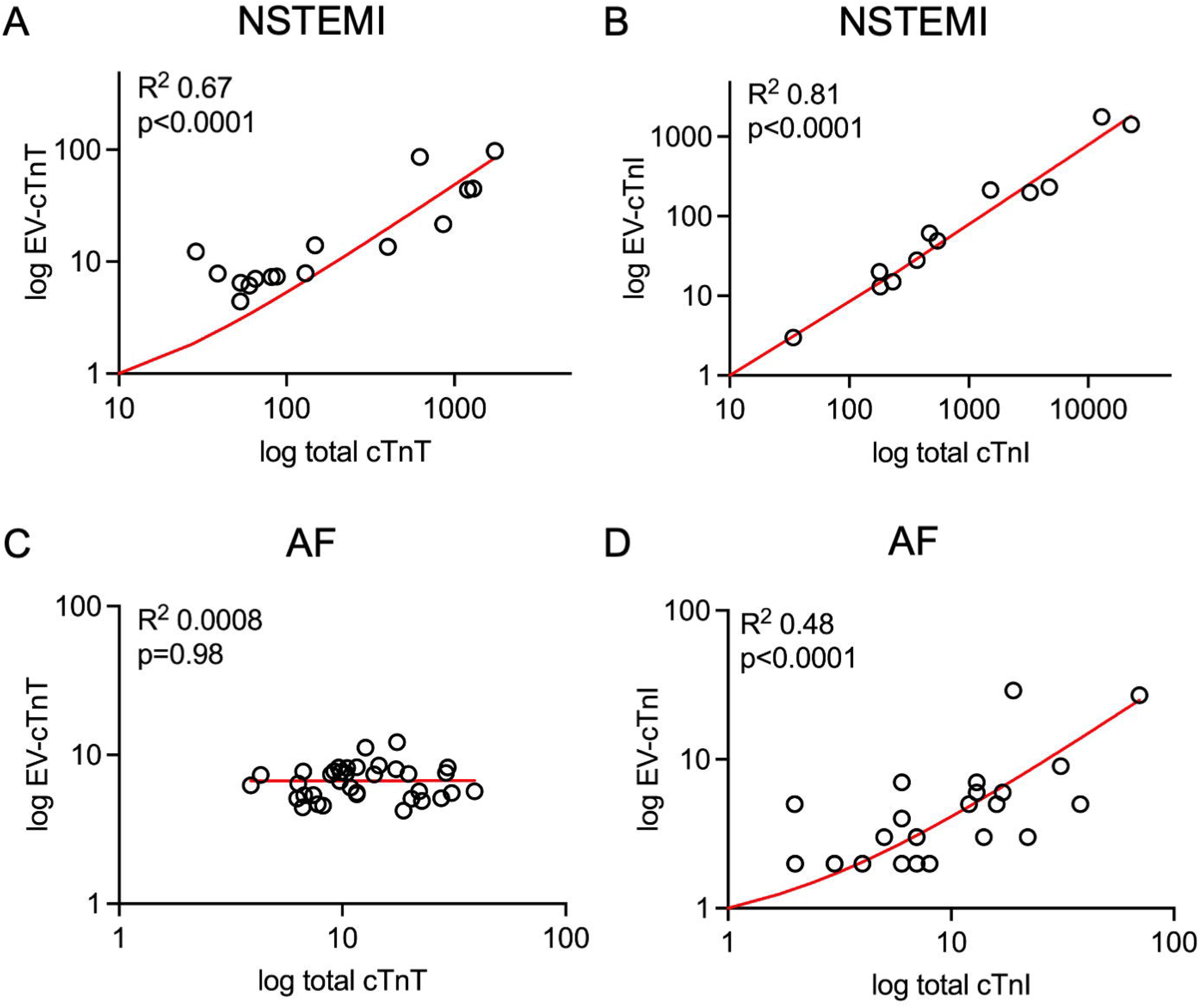
Correlative analysis of EV-bound fraction and total cTnT and cTnI in myocardial ischemia and strain states points towards disease-specific troponin release kinetics. Simple linear regression of total vs. EV-bound fraction of **A** cTnT (n=16) and **B** cTnI (n=12) in the NSTEMI cohort, and of **C** cTnT (n=37) and **D** cTnI (n=32) in the atrial fibrillation (AF) cohort.

## Discussion

The present study provides further evidence in the emerging field of cardiac troponins enclosed within the EV compartment in circulation, advancing the role of EV-bound cardiac troponin T and I as potential biomarkers for cardiovascular disease. Herein, we are, to our knowledge, first in describing disease-specific EV- and non-EV-troponin compartmental signatures, allowing to robustly discriminate temporary myocardial strain secondary to tachyarrhythmia from type I myocardial infarction via a single blood sample. Furthermore, present data indicates a potential role for blood EV-troponin signatures to distinguish firstly manifesting atrial fibrillation or flutter from a recurrent episode, thus potentially simplifying downstream therapeutic decision-making. To this end, EV-troponin signatures and EV-troponin fragmental analysis may be the long-awaited tool to enhance diagnostic specificity of current state-of-the-art cardiac troponin assays, with the potential to save clinical resources presently exhausted in the diagnostic workup of unclear troponin elevation. The concept of an active secretory pathway of cardiac troponins into the bloodstream as part of the physiological stress response of the myocardium may further provide a missing link to understanding the mechanism of troponin release in the absence of necrosis. In various diseases associated with myocardial strain, ranging from arrhythmias, congestive heart failure, inflammatory heart disease to pulmonary embolism, a moderate blood troponin elevation with variable resolution over time and oftentimes no morphological evidence of myocardial scarring is described in present literature. These observations raise the hypothesis of an alternative pathway of troponin extradition from diseased but viable cardiomyocytes. Enrichment of cardiac troponins in EV released by stressed cardiomyocytes into circulation, which are then degraded over time by blood proteases found to be elevated in physiological organismal stress responses (27), may provide a plausible mechanistic explanation for increased troponin levels in the setting of temporary myocardial strain.

In overt myocardial ischemia and myocardial hypoxic stress, increased systemic EV release into circulation concomitant with a global elevation of cTnT enriched in EV can be derived from *in vitro* and murine *ex vivo* experiments. Combined with the release of structural troponin T from apoptotic cardiomyocytes into circulation, these observations postulate an accelerated release of EV from diseased cardiomyocytes that may precede or accompany irreversible myocardial necrosis. Patient data from two independent cohorts suffering from type I myocardial infarction (NSTEMI) requiring recanalization of at least one critical stenosis (>70%) demonstrate the relative predominance of unbound cardiac troponins in blood in the setting of necrosis, with a dramatic shift of >85% of total cTnT/cTnI compartmentalized in the free-plasma fraction. In our in-vitro model, global hypoxic stress induced cysteine protease-dependent cytosolic fragmentation of troponin T in cardiomyocytes without enhanced apoptosis. In the context of arrhythmia-induced temporary myocardial strain, a moderate increase in plasma cTnT levels coincided with a ∼1.9-fold increase in systemic circulating EV concentration. These data suggest an EV predominance of circulating troponin in myocardial strain distinct from compartmental troponin signatures observed in myocardial necrosis, which was confirmed in our fractional troponin analysis demonstrating ∼55% of plasma cTnT and ∼63% of plasma cTnI compartmentalized in EV. These observations give rise to a number of hypotheses on circulating troponin kinetics: The heterogeneous disease entities commonly termed acute myocardial damage, type II myocardial injury or myocardial infarction/ischemia with non-occlusive coronary arteries (MINOCA/INOCA) may represent a conjoined pathway triggered by different myocardial stressors. Herein, the stressed cardiomyocyte secretes EV enriched with cardiac troponins into the bloodstream as an initial response, which are postulated to release their molecular content through physiological degradatory processes, causing the observed moderate and delayed plasma troponin elevation. As myocardial strain persists over time, persistent EV secretion presumably combined with a small extent of cardiomyocyte apoptosis below the detection threshold of cardiac imaging techniques may entertain said troponin elevation. Furthermore, in cases of persistent and/or frequently recurrent myocardial strain as in atrial fibrillation or flutter, myocardial remodeling processes may alleviate the strain response over time to achieve a homeostasis-like steady state. This theory may explain the cTnI compartmental signatures observed in recurrent, but not in firstly diagnosed AF.

In summary, present evidence delineates relative EV-bound cardiac troponin fraction and troponin compartmental distribution patterns in circulation as a novel biomarker in cardiovascular disease with potential disease entity and disease stage-specific signatures.

Further studies may identify EV-troponin signatures specific to other causes of myocardial strain commonly associated with transient troponin elevation or type II myocardial injury such as inflammatory heart disease, acute or chronic congestive heart failure, pulmonary embolism, or drug-induced cardiotoxicity. Moreover, mechanistic studies are required to address the origin of free-plasma cardiac troponin as well as the specific pathway of troponin disintegration and subsequent EV enrichment in cardiomyocytes in these disease contexts. Another intriguing question is whether structural differences in cardiac troponins detected in EV and non-EV fractions may hold additive diagnostic value. The observed blood troponin compartmental signatures may further address currently open questions with regard to disease context-specific prognostic implications or specific therapeutic strategies.

### Limitations

The present study is primarily of hypothesis-generating nature and its derivative results are limited by sample size and biological variation. Advanced studies will need to verify the observed blood troponin signatures on a larger scale and investigate their translation to other disease contexts. At present, identification and quantification of relative EV-troponin fraction is time and cost-intensive, and technical automation will be necessary to establish this methodology in clinical routine, especially to accommodate the time constraints of an emergency department setting.

### Clinical perspectives

The specific EV-troponin compartmental signatures identified herein hold the potential to enhance diagnostic specificity with regard to cardiovascular disease entity, disease stage with a particular focus on early diagnosis of myocardial disease preceding irreversible necrosis, as well as prognostication. A further elaboration of this novel biomarker may promote diagnostic accuracy and aide early-stage diagnostic and/or therapeutic interventions derived from a single blood sample, initiated as early as upon first symptomatic disease manifestation.

### Competency in medical knowledge

Gaining a deeper mechanistic understanding of the dynamics of EV release and troponin distribution in circulation may reshape our present knowledge of myocardial pathophysiologies, with the potential to redefine currently established disease entities based on different underlying pathomechanisms.

### Translational outlook

This study may open up a novel perspective in translational research with a focus on cardiac troponins and their specific compartmental distribution in blood in various disease contexts. Further exploration of the pathological pathways conducive to different troponin release mechanisms may aide in the timely diagnosis of myocardial disease, thus guiding resource-sparing diagnostic measures and disease-specific therapies.

## Data Availability

All data produced in the present study are available upon reasonable request to the authors.

## Acknowledgments

The authors would like to express their gratitude to Amelie Werner and Constanze Winkler for their invaluable technical assistance throughout this project.

## Relationship with Industry Declaration

The authors have received project-related funds and materials from Roche Diagnostics.

## Non-Standard Abbreviations and Acronyms

EV: extracellular vesicles
cTnT/cTnI: cardiac troponin T/I
hs-cTnT/I: cardiac troponin T/I quantified by high-sensitive assay
NSTEMI: non-ST segment elevation myocardial infarction
AF: atrial fibrillation
AFlutt: atrial flutter
PB: peripheral blood

---

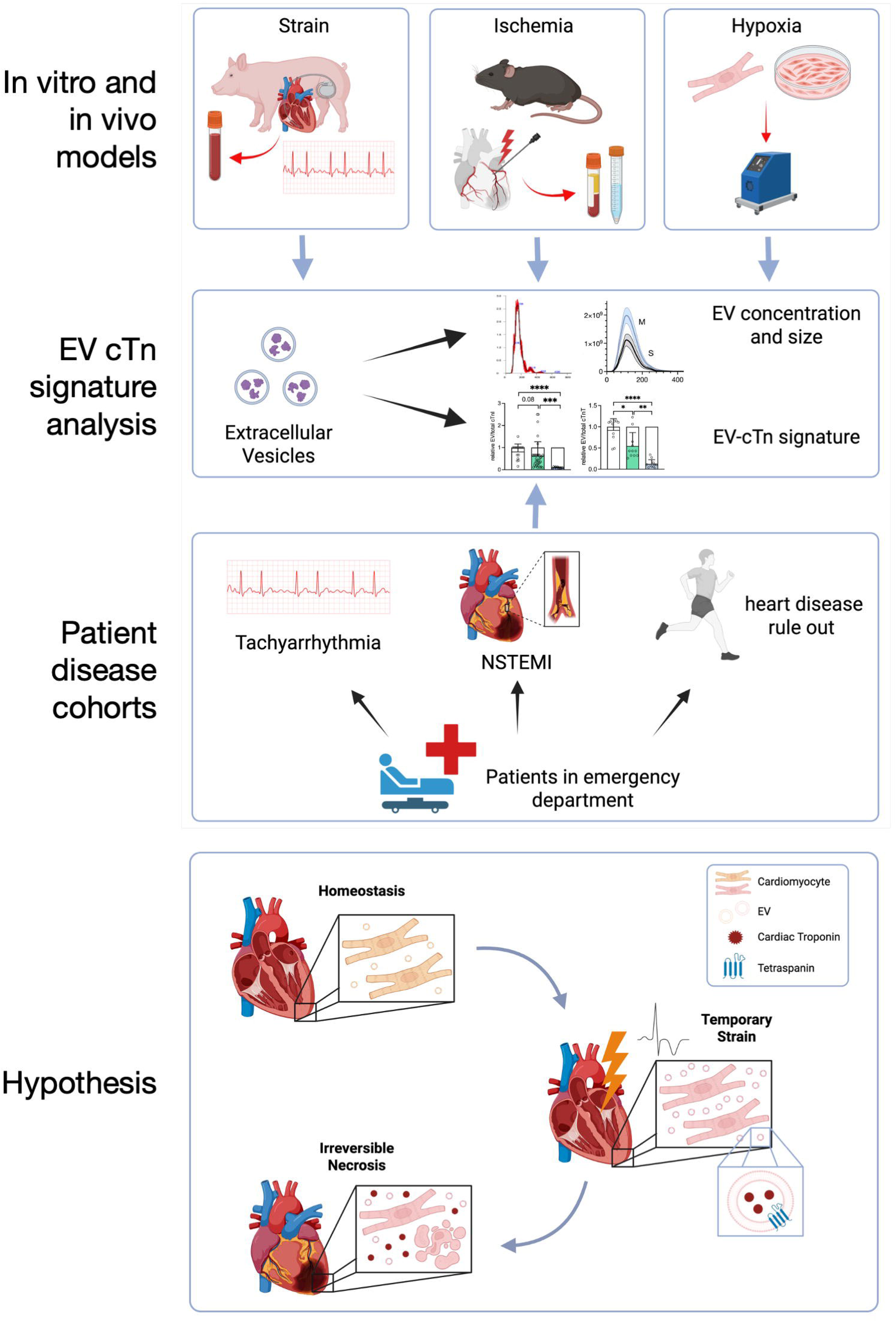
Visual Abstract: Extracellular vesicles extracted from cardiomyocytes under hypoxia in vitro, mice following minimal-invasive myocardial infarction and pigs after induction of sustained tachyarrhythmia are analyzed in terms of EV concentration, size distribution and relative EV-bound cardiac troponin (cTn) fraction. In patients presenting to the emergency department with tachyarrhythmia, NSTEMI or individuals in whom heart disease can be ruled out, blood EV are isolated and analyzed accordingly. This study hypothesizes that cTn-containing EV are constitutively secreted by cardiomyocytes under homeostasis, and temporary myocardial strain states induce EV secretion, causing cTn release in the bloodstream in an EV-predominant manner. In irreversible myocardial necrosis, cTn is released into circulation in its unbound form. Relative EV-bound to free-plasma troponin fractions can increase diagnostic specificity for different myocardial diseases.

