## Supplemental Figures for "A circulating extracellular vesicle-bound fraction of cardiac troponin discriminates myocardial homeostasis and disease states"

### Slide 1
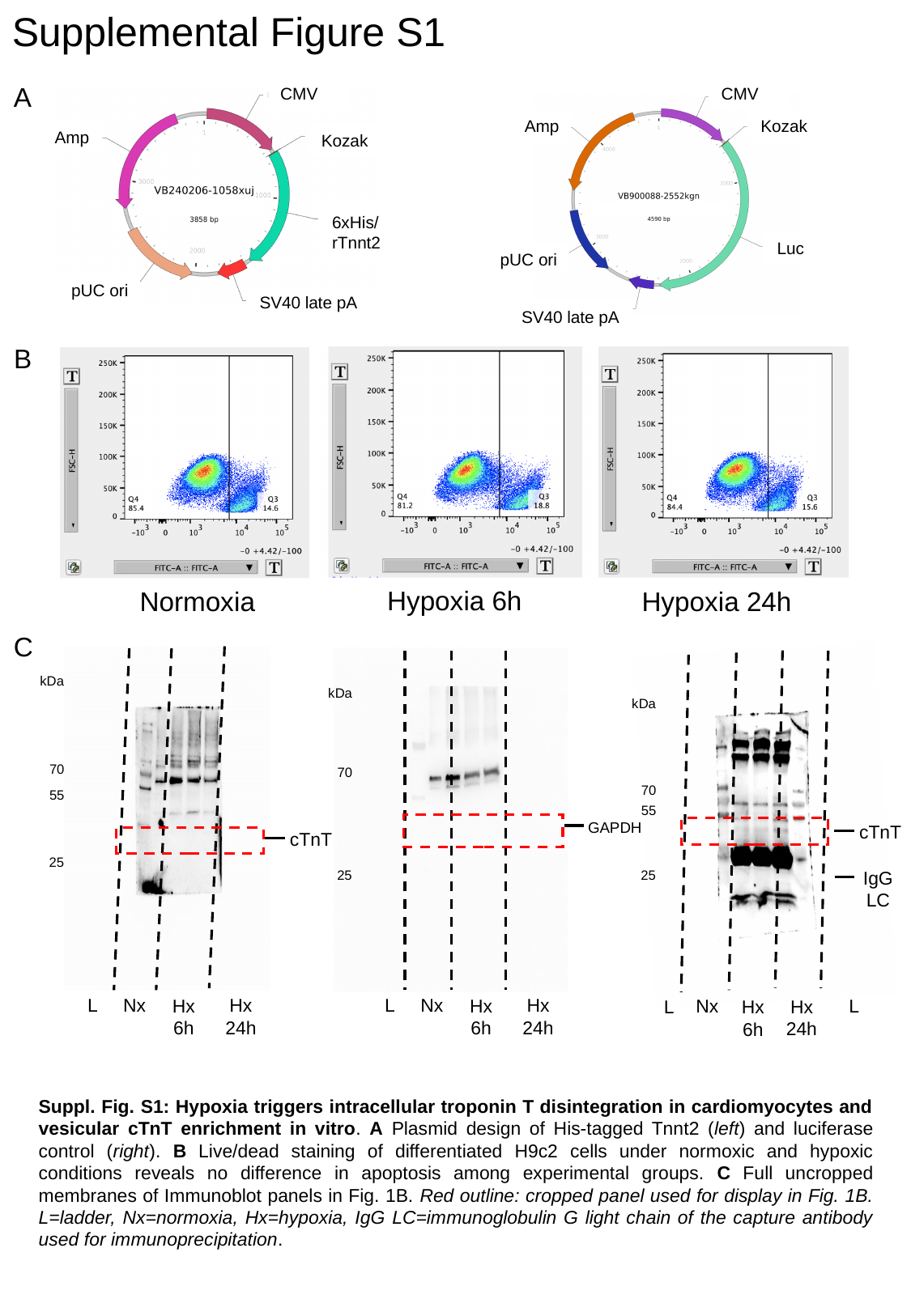

Supplemental Figure S1
A
CMV
CMV
Kozak
Amp
Amp
Kozak
6xHis/rTnnt2
Luc
pUC ori
pUC ori
SV40 late pA
SV40 late pA
B
Hypoxia 6h
Normoxia
Hypoxia 24h
C
kDa
kDa
kDa
70
70
70
55
55
GAPDH
cTnT
cTnT
25
25
IgG
LC
25
Nx
Nx
L
L
Hx
24h
Hx
24h
Hx
6h
Hx
6h
L
Nx
L
Hx
24h
Hx
6h
Suppl. Fig. S1: Hypoxia triggers intracellular troponin T disintegration in cardiomyocytes and vesicular cTnT enrichment in vitro. A Plasmid design of His-tagged Tnnt2 (left) and luciferase control (right). B Live/dead staining of differentiated H9c2 cells under normoxic and hypoxic conditions reveals no difference in apoptosis among experimental groups. C Full uncropped membranes of Immunoblot panels in Fig. 1B. Red outline: cropped panel used for display in Fig. 1B. L=ladder, Nx=normoxia, Hx=hypoxia, IgG LC=immunoglobulin G light chain of the capture antibody used for immunoprecipitation.

### Slide 2
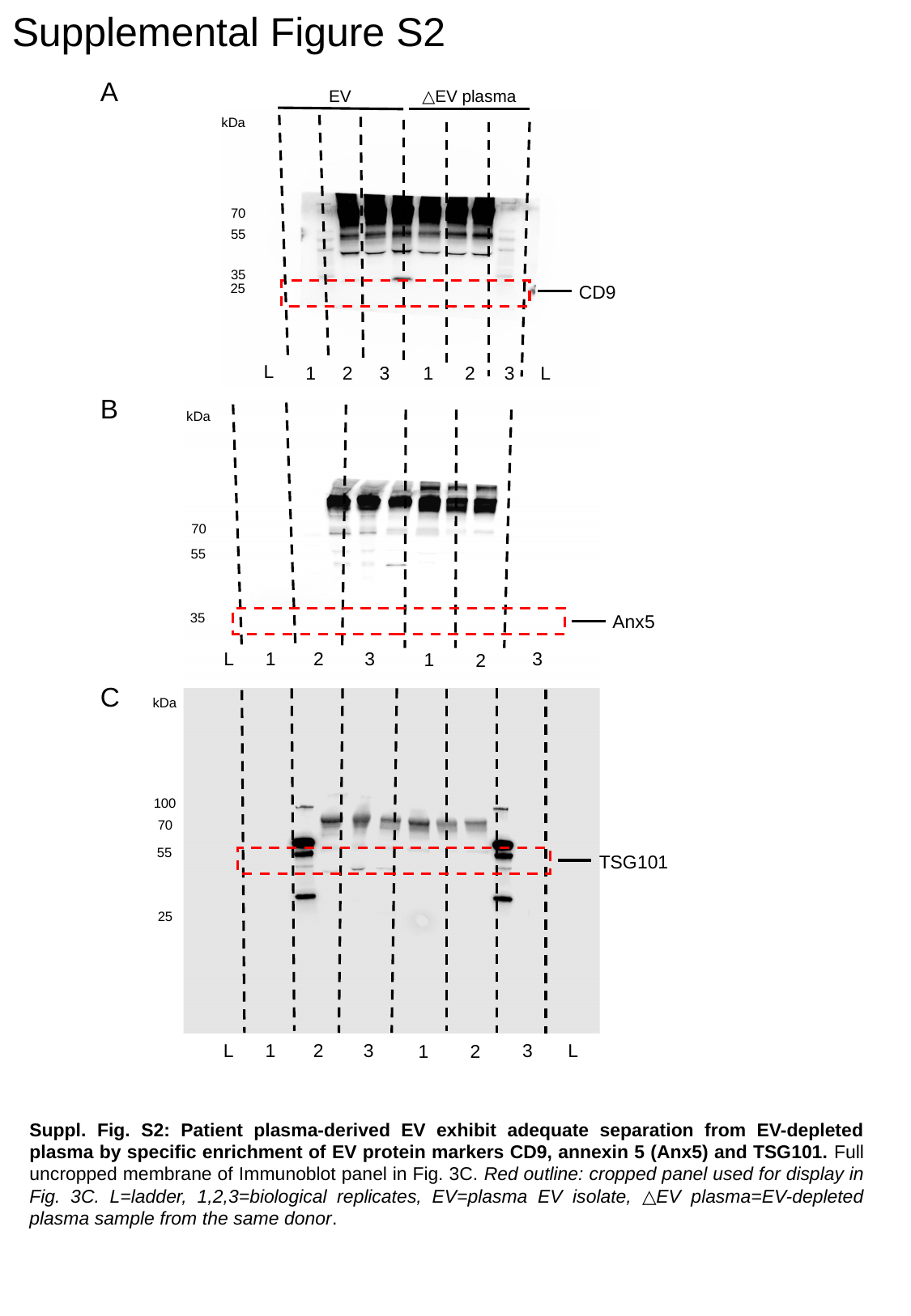

Supplemental Figure S2
A
EV
△EV plasma
kDa
70
55
35
25
CD9
L
1
2
3
1
2
3
L
B
kDa
70
55
35
Anx5
L
1
2
3
3
1
2
C
kDa
100
70
55
TSG101
25
L
L
1
2
3
3
1
2
Suppl. Fig. S2: Patient plasma-derived EV exhibit adequate separation from EV-depleted plasma by specific enrichment of EV protein markers CD9, annexin 5 (Anx5) and TSG101. Full uncropped membrane of Immunoblot panel in Fig. 3C. Red outline: cropped panel used for display in Fig. 3C. L=ladder, 1,2,3=biological replicates, EV=plasma EV isolate, △EV plasma=EV-depleted plasma sample from the same donor.

### Slide 3
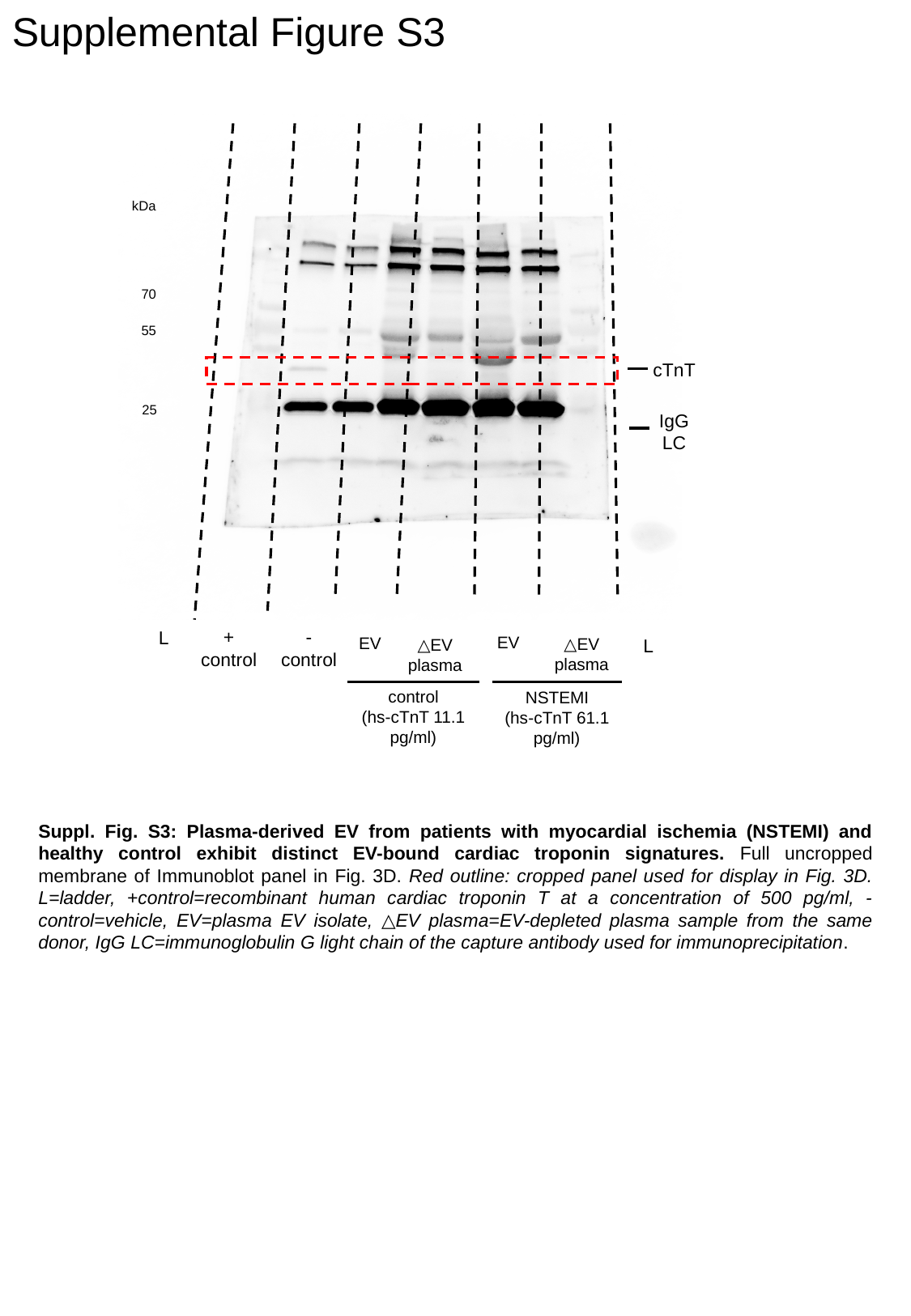

Supplemental Figure S3
kDa
70
55
cTnT
25
IgG
LC
-
control
L
+
control
EV
EV
△EV
plasma
L
△EV
plasma
control
(hs-cTnT 11.1 pg/ml)
NSTEMI
(hs-cTnT 61.1 pg/ml)
Suppl. Fig. S3: Plasma-derived EV from patients with myocardial ischemia (NSTEMI) and healthy control exhibit distinct EV-bound cardiac troponin signatures. Full uncropped membrane of Immunoblot panel in Fig. 3D. Red outline: cropped panel used for display in Fig. 3D. L=ladder, +control=recombinant human cardiac troponin T at a concentration of 500 pg/ml, -control=vehicle, EV=plasma EV isolate, △EV plasma=EV-depleted plasma sample from the same donor, IgG LC=immunoglobulin G light chain of the capture antibody used for immunoprecipitation.
