## Supplementary material for "A circulating extracellular vesicle-bound fraction of cardiac troponin discriminates myocardial homeostasis and disease states": Tables 1-2

|  | **Control**  **(n=10)** | **Non-STEMI**  **(n=11)** | **AFib/AFlut**  **(n=11)** | **p value** |
| --- | --- | --- | --- | --- |
| **Age (yrs)** | 47.2(±18.5) | 67.5(±7.5) | 70.4(±6.4) | <0.001 |
| **Male gender** | 6/10 | 10/11 | 9/11 | 0.23 |
| **BMI (kg/m²)** | 26.3(±4.4) | 27.4(±5) | 27.7(±3.3) | 0.71 |
| **Systolic BP (mmHg)** | 152.7(±16.4) | 157.4(±16.7) | 146.5(±18.8) | 0.35 |
| **Heart rate (*min^-1^)** | 98.3(±25) | 76.2(±12.1) | 115.8(±20.5) | <0.001 |
| **Median days since onset of symptoms** | 1.5 [0.3-5.8] | 1.0 [0.2-10.5] | 0.5 [0.25-2.4] | 0.89 |
| **Serum hs-cTnT (pg/ml)** | 6.1(±2.3) | 226.8(±191.5) | 12.6(±5) | 0.032 |
| **eGFR (ml/min)** | 99.6(±18.9) | 73.5(±26.7) | 79.9(±14.6) | <0.001 |

**Table 1: Patient demographics of pilot cohort**

**Table 2: Patient demographics of validation cohort**

|  | **Control**  **(n=23)** | **Non-STEMI**  **(n=12)** | **AFib/Flut**  **(n=36)** | **p value** |
| --- | --- | --- | --- | --- |
| **Age (yrs)** | 37.1(±11.6) | 67.7(±10.8) | 66.9(±13.2) | <0.001 |
| **Male gender** | 14/23 | 8/12 | 25/36 | 0.79 |
| **BMI (kg/m²)** | 25.7(±3.8) | 27(±8.1) | 28.1(±5) | 0.26 |
| **Systolic BP (mmHg)** | 129.4(±11.9) | 146.8(±29.6) | 138.4(±17.7) | 0.016 |
| **Heart rate (*min^-1^)** | 76.2(±12.6) | 77.8(±16.7) | 111.2(±24.2) | <0.001 |
| **Median days since onset of symptoms** | 1.5 [0.7-4.8] | 1.0 [0.7-3.5] | 2.0 [0.7-4.0] | 0.94 |
| **Serum hs-cTnT (pg/ml)** | 5.5(±3.1) | 305.1(±389.5) | 14.7(±8.6) | <0.001 |
| **Serum hs-cTnI (ng/L)** | 4.4(±6.9) | 3633.2(±6115.1) | 13.2(±14.3) | <0.001 |
| **eGFR (ml/min)** | 106.5(±15.6) | 80.6(±23.7) | 81.7(±16) | <0.001 |
